# Mixed-methods assessment of engagement with a digital intervention: the Wrapped feasibility Randomised Controlled Trial

**DOI:** 10.1101/2025.09.16.25335892

**Authors:** Lauren Schumacher, Kayleigh Kwah, Rik Crutzen, Katherine Brown, Stephen Bremner, Louise J Jackson, Katie Newby

**Affiliations:** Public Health and Applied Behaviour Change (PHAB) Lab, University of Hertfordshire, Hatfield, UK; Department of Health Promotion, Care and Public Health Research Institute (CAPHRI), Maastricht University, Maastricht, the Netherlands; Department of Primary Care and Public Health, Brighton and Sussex Medical School, Brighton, UK; Department of Applied Health Sciences, College of Medicine and Health,, University of Birmingham, Birmingham, UK

## Abstract

Digital health behaviour change interventions can face the challenge of low participant engagement, which can limit intervention effectiveness. Mixed methods approaches to understanding engagement, which capture the online and offline behaviours of participants, as well as the cognitive and affective aspects of engagement, are infrequently reported. The aim of this study was to explore these aspects of engagement for a digital intervention (Wrapped) to enable its optimisation ahead of testing in a randomised controlled trial (RCT). Wrapped is a digital intervention that aims to increase correct and consistent condom use, thereby decreasing the incidence of sexually transmitted infections among young people aged 16-24 years. Analytics data and website user history data were combined with data from surveys and qualitative interviews. Together this data was examined to assess the behavioural, cognitive, and affective aspects of engagement with the intervention. Results showed that participants experienced few barriers during the registration process, but that thetailoring questions used to assign content to individual users may not have been working as intended. Pre-determined intervention goals were as follows: ordering Sample Pack 48 (60.8%), using Condom Ordering Service 37 (52.9%), ordering Condom Carrier 31 (49.2%), watching Condom Demo video 7 (10%), watching Discussing Condoms video 4 (6.8%), and watching Real Life video 9 (13.6%). Participants described their use and enjoyment of the products they ordered; notably the condom carrier was less well liked and used. Participants reported not engaging with the video components, either because they were unaware that they existed or because they expected to find watching them to feel awkward. This study demonstrates that taking a mixed-methods approach to studying engagement provides a more complete understanding of where and how digital interventions need to be optimised than using single methods in isolation; this in turn is likely to lead to more effective interventions.

**Author Summary:** For a digital health behaviour change intervention to be effective, participants must engage with the intervention. Unfortunately, low engagement is a challenge for many digital interventions. Few studies combine quantitative and qualitative data to capture the behavioural (both online and offline), cognitive, and affective aspects of engagement. The aim of the present study was to explore how a digital intervention (Wrapped) was used and to further understand the engagement experience to provide insights to guide future intervention optimisation. Wrapped is a digital intervention for 16-24 year olds that aims to increase correct and consistent condom use, thereby decreasing sexually transmitted infections (STIs). Analytics data and website user history data were combined with data from surveys and qualitative interviews. Results indicated that participants experienced few barriers in registering for the intervention or in ordering free products (e.g. a condom sample pack and carrier); participants also reported that they used and enjoyed the products that they received. Most participants did not engage with the video content, either because they expected to find watching them awkward or because they did not know they existed. Mixed-methods research better supports researchers to make decisions about the optimisation of digital interventions than using single methods alone.

## Introduction

Digital health behaviour change interventions offer users the flexibility and ease to engage at a time and place that is convenient to them (1) and the privacy to engage with an intervention that might otherwise be associated with feelings of embarrassment or stigma (2). They also have good potential reach; internet use is now near-universal amongst UK adults (3), and digital delivery can remove barriers to access typically associated with in-person delivery (1). In addition to the benefits for users, the potential to deliver digital interventions with high fidelity (4), likely at lower-cost than is the case for face-to-face interventions, also makes them attractive to developers. For researchers wishing to evaluate digital behaviour change interventions, internet-based trials can however present challenges, most notably low participant engagement with intervention content which can negatively impact intervention effectiveness (5). Intervention effectiveness is understood to be mediated by engagement (6–8); if a user does not engage with the content then they do not stand to benefit, and any positive effects which might have eventuated will go unobserved. Intervention engagement in a trial context is therefore a marker of successful trial delivery (6). As such, researchers preparing to test digital interventions are interested in measuring and understanding intervention engagement before the research begins, so that any barriers can be addressed and the potential for engagement optimised.

Although it is generally accepted within the field of digital interventions that engagement is important, arriving at a shared understanding of what engagement is has proved challenging (9). Previous definitions of engagement have focused almost solely on the behavioural aspects of engagement, that is, how an intervention is used (9). In the case of digital interventions this has typically been measured using data analytics, which records, for example, repeat visits to an application, the duration of each visit, which pages and content are accessed, etc. (8). Collecting analytics data is an attractive means of measuring engagement with digital interventions as it places no reporting burden on participants, has limited impact on their intervention experience and, with careful pre-planning of what data to track, can easily inform how they are using the intervention (7).

Defining intervention usage solely in analytics-terms can however be limiting as it ignores any offline activities associated with the intervention, such as behaviours or actions performed “in real life” (6). Analytics provide answers to questions about how participants engage with digital interventions themselves, but not about any activity in the offline world that more directly brings about the desired intervention outcome (e.g., taking a walk, making dietary changes, using a condom, etc.) (10). Although analytics data tends to be collected and analysed most frequently for digital interventions, other types of data, such as that generated through qualitative methods (e.g. focus groups, interviews), self-report questionnaires, Ecological Momentary Assessment (EMA), and sensors (e.g. accelerometers), can yield important insights into offline engagement (7).

Examining engagement purely through the analytics lens also neglects the cognitive and affective aspects of engagement with digital interventions (9). Cognitive aspects of engagement can be thought of as interest, attention and focus (8–10). Affective aspects, on the other hand, refer to both the positive and negative emotional experiences related to engagement, which encompass satisfaction or enjoyment, a sense of community with the intervention and other users, and motivation or desire to continue using the intervention (6,10). Qualitative analyses, via interviews, focus groups, and think aloud studies (8), can identify these cognitive and affective aspects of engagement. These are considered as important to consider as the behavioural aspects, especially as interventions are thought to be more effective when participants enjoy engaging with them (8), and highlight why participants engage in the first place and in what context (11).

Where analytics can answer questions about how participants engage with online aspects of digital interventions, other research methods can answer questions about how they engage with offline aspects, why they engage, and in what context (6). To address these questions, mixed methods analyses are needed (6–8,12). This paper presents a holistic, mixed methods approach to measuring engagement for the digital intervention Wrapped. It draws upon previous definitions of engagement from both Perski et al. (8) and Kelders et al. (9) exploring the amount the intervention was used online and offline (how much, how often, why, in what context, and to what extent), and the cognitive and affective experience of participants when using the intervention (8,9). The aim of this study was to explore the extent to which a digital intervention (Wrapped) was used during a feasibility randomised controlled trial (fRCT) and to further understand the engagement experience to inform optimisation of the intervention for future testing in a definitive randomised controlled trial (RCT).

## Methods

### The Wrapped intervention

Wrapped is a digital intervention that aims to increase correct, consistent condom use, thereby decreasing sexually transmitted infections (STIs) among young people aged 16-24 years. It was co-created in 2016-17 by researchers, professional stakeholders, and young people. Wrapped is a multi-component website that provides access to orderable products and video material. Content is tailored to each user’s self-identified barriers to condom use. Users are given access to between one and six components. These components are allocated according to their answers to tailoring items presented to them when they first access the site. The six components and the relevant tailoring items are described in table 1 below. To be allocated any one component, users had to agree to one or more of the specified questions. Due to the sexually explicit nature of component six, only users aged 18 years or older were allocated this content. For more details on the Wrapped intervention and its development, see Newby et al. 2019 (13).

**Table 1.** Tailoring of components for Wrapped intervention.

| Component | Description | Tailoring items* |
| --- | --- | --- |
| 1. Sample Pack | A single order of a sample pack of 12 different types of condoms, 2 sachets of lubricant, and instructions on how to use condoms. The sample pack was posted in discreet packaging. Participants could order only once. | N/A<br>This component was offered to all participants regardless of responses to the tailoring items. This ensured that all users received some content. |
| 2. Condom Ordering Service | A free monthly ordering service; participants were sent monthly reminders to place an order for condoms (of a single type) in bundles of either 6 or 12 and up to 3 sachets of lubricant. Participants could also leave reviews for condoms on the website. A maximum of 12 orders could be placed. | "I can't always get the type of condoms I want."<br>"I find condoms expensive to buy."<br>"I find buying condoms embarrassing." |
| 3. Condom Carrier | A single order of a keychain wallet with a discreet compartment for a condom; two designs to choose from. | "I don't always have a condom on me when it's needed." |
| 4. Condom Demo Video | A single video featuring several groups of young people offering instructions on how to correctly use condoms, with tips on how to make using condoms pleasurable for both partners. | "I'm not always able to put a condom on with confidence and ease."<br>"I find using condoms interrupts the flow of sex."<br>"Condoms make sex less enjoyable or pleasurable for me."<br>"Condoms make sex less enjoyable or pleasurable for the person I'm with." |
| 5. Discussing Condoms Videos | Seven individual videos featuring groups of young people talking about how to discuss using condoms with partners. | "I find it awkward or difficult letting someone know that I want to use condoms."<br>"I find using condoms interrupts the flow of sex." |
| 6. Real Life Videos | Three videos featuring real couples (two heterosexual and one gay) talking about and demonstrating actual condom use during sex. No footage of genitals or penetration.<br>*These videos were only shown to participants aged 18 years or over.* | "I find using condoms a turn-off."<br>"I find using condoms interrupts the flow of sex."<br>"Condoms makes sex less enjoyable or pleasurable for me."<br>"Condoms makes sex less enjoyable or pleasurable for the person I'm with." |

\*Participants are assigned the component if they answer yes to any of the items in associated cell of this column

### The Wrapped feasibility randomised controlled trial

Data was collected in a feasibility randomised controlled trial (fRCT) between March 2021 and October 2022 to test whether and how it would be possible to run a full trial of the Wrapped intervention. The target sample size of 230 participants was based on the recommended number of participants required to estimate design parameters for a full trial with good precision, inflated to allow for expected drop-out (14). The target sample of 230 16-24 year olds was recruited from one of two websites offering sexually transmitted infection (STI) self-sampling (freetest.me and SH.UK; both owned by Preventx Ltd). Users viewed an advert for the study upon completing their order for a free STI self-sampling test kit, and interested users followed a link to the study participant information sheet and consent form on REDCap (15), a secure data capture and management platform. Following completion of a baseline survey on REDCap, participants, blinded to allocation, were randomised to receive the Wrapped intervention or usual care information (basic information on STIs and condoms use) presented via a control website. Participants who accessed either website received a £5 shopping voucher. Those who did not access the website were sent a reminder two weeks later via email. Explicit consent was obtained from participants for collecting analytics data on their use of either the Wrapped or the control website. Participants completed a further three surveys via REDCap at months 3, 6, and 12, as well as taking two further STI self-samples at months 3 and 12. For more details on the Wrapped fRCT see (16).

For the purposes of this paper, we only report on data collected about participants randomised to the intervention condition (n=115), as only these participants received the intervention content we were seeking to optimise.

### Measuring online engagement

#### Predetermined intervention goals

Ahead of data collection, a set of intervention goals that would provide a binary measure of participant engagement with the registration process and each of the intervention components was determined (see table 2). A combination of analytics data and user order histories were used to determine if an intervention goal was achieved.

**Table 2.** Predetermined goals for six Wrapped intervention components.

| Component | Predetermined intervention goal |
| --- | --- |
| Registration | Complete website registration, answer all tailoring items, and reach the homepage |
| Sample Pack | Order sample pack |
| Condom Ordering Service | Place at least one order for condoms |
| Condom Carrier | Order condom carrier |
| Condom Demo Video | Watch condom demo video (any duration) |
| Discussing Condoms Videos | Watch a minimum of one of the discussing condoms videos (any duration) |
| Real Life Videos | Watch a minimum of one of the real life videos (any duration) |

#### Analytics data

Matomo (17), a free opensource web analytics application, was used to collect the analytics data. To enable individual-level analytics data to be collected, Matomo was set up to capture each user’s participant ID (PID) and associate it with data collected on their use of the site each time they accessed it. This was achieved by providing users with a URL that had their PID appended to the end. The PID was captured by the site during the registration process and then associated with each individual’s user account. At subsequent visits, users were required to log-in with their account details, and thus data was automatically associated with their PID. Each page was tracked by Matomo, with the software recording for each PID, if and how the individual engaged with the content. The following information was recorded for each participant:

1. If registration was completed.
2. If the tailoring items were completed.
3. Which components were allocated.
4. Which component pages were accessed (how many times and how long each visit was).
5. If a Sample Pack was ordered.
6. If a first order was placed for the Condom Ordering Service.
7. If subsequent orders (and how many) were placed for the Condom Ordering Service.
8. If a Condom Carrier was ordered.
9. If the Condom Demo Video was watched and if so, for how long.
10. If any of the Discussing Condoms Videos were watched and if so, which ones and for how long.
11. If any of the Real Life Videos were watched and if so, which ones and for how long.

Data was also captured on whether the pages where orders were placed were visited, and if users began ordering but did not complete the process. To enable this individual-level data to be collected, users were required to register at first access to the website and to sign-in on subsequent occasions. On completion of the fRCT, participants’ analytics data were downloaded from Matomo in Microsoft Excel (18) format; each row representing an individual participant with the columns displaying the analytics data. The Excel file was imported into SPSS (19) for analysis.

#### User history data

User order histories were recorded within the Wrapped website, accessible via the admin portal. Order histories captured information on whether the condom sample pack, the condom carrier, and/or any condom and lubricant bundles were ordered, and in the case of the latter, how many orders were placed. Data was also captured on whether the pages where orders were placed were visited, and if users began ordering but did not complete the process. To enable this individual-level data to be collected, users were required to register at first access to the website and to sign-in on subsequent occasions. On completion of fRCT data collection, order histories along with participant IDs were downloaded from the website in Microsoft Excel format.

The analytics and order history data were combined into a single Excel file; each row representing an individual participant with the columns displaying their order history. The Excel file was imported into the existing SPSS file for analysis.

#### Survey data

Information about online engagement with Wrapped was also collected by survey. This was used for two purposes. Firstly, survey data was used to assess the accuracy by which tailoring items allocated intervention content to users. As described above, tailoring items were used to identify each participant’s barriers to condom use. Ten items in the baseline survey measured behavioural determinants of condom use that were aligned to these barriers (see Supplementary File 1 for the questions and alignment to the barriers). These items were used to cross-check allocation, first by assigning components based on the survey questions, and then by comparing the proportion of participants allocated via the two different approaches using cross-tabulation. See Supplementary File 1 for more detail on how this analysis was conducted. Secondly, survey questions were used to assess perceptions of the usefulness of the website and the quality of design. Relevant questions to measure this were included in the first follow-up survey at 3 months (see Supplementary File 2 for the questions used). Survey data was downloaded from REDCap as an Excel file and imported into the existing SPSS file for analysis.

### Measuring offline engagement

#### Survey data

The final follow-up survey, sent to participants 12 months from baseline, included questions used to measure offline engagement with Wrapped materials. These were used to assess whether ordered items were used, whether accompanying information was read, and to gather some initial feedback (see Supplementary File 2 for the questions used). At the end of fRCT data collection, this survey data was downloaded from REDCap in Microsoft Excel format and imported into the existing SPSS datafile for analysis.

#### Qualitative data

Participants were asked in the baseline survey, if they would be interested in taking part in a semi-structured telephone interview, the purpose of which, in part, was to gather feedback on the intervention materials (for details on other objectives of the qualitative interviews and results, see Newby et al (20). A total of 89 (38.7%) individuals expressed an interest in this, with 30 (33.7%) going on to participate. Data was collected at three timepoints, with ten participants providing data at each (months 3, 6, and 12). Half of the participants were those randomised to the intervention condition, and half to the control. For the purposes of this paper, only data provided by participants in the intervention condition is reported (n=15).

To obtain a representative sample, participants were selected based on their level of engagement, and their demographic characteristics such as age, gender, ethnicity, sexual identity, and level of deprivation. One participant asked whether the interview questions and their responses could be in written form to accommodate their access needs. Accordingly, for this one participant, responses were provided by secure online survey software instead of by interview. Interviews ranged from 9.1 to 49.5 minutes, averaging at 24.8 minutes. Interviews were audio recorded and transcribed verbatim. Written responses from the individual given the survey were downloaded and treated as a transcript.

Framework analysis was used to analyse the transcripts; this approach was chosen as it allows for themes to be examined across participants while preserving the individual context of each participant (21). Two researchers (KK and LS) initially coded four transcripts independently using a preliminary agreed framework, discussing required changes based on codes that did not fit. The framework was then finalised and used by KK to code the remaining transcripts. NVivo (22) was used to support coding. Microsoft Excel was used to build a framework matrix that included a summary of the data and illustrative quotes for each code.

### User experience (UX) review

A user experience (UX) expert was commissioned to provide an independent, specialist usability review of the Wrapped intervention. This review was conducted without knowledge of the fRCT findings relating to user engagement as reported in this paper. The website was evaluated against established and recognised usability principles (23). The information architecture, or the “process of designing, implementing, and evaluating information spaces that are humanly and socially acceptable to their intended stakeholders”(24), was also examined. The findings of this review are included in this paper, and discussed alongside the fRCT data, as they add an extra layer of insight into user engagement.

### Ethical approval

The study was registered as International Standard Randomised Controlled Trial Number (ISRCTN) 17478654 (25). Ethical approval was granted by NHS East Midlands - Leicester Central Research Ethics Committee (reference 20/EM/0275). The fRCT itself was registered as International Standard Randomised Controlled Trial Number (ISRCTN) 17478654. Formal consent was obtained for all participants via REDCap.

## Results

### Accessing and Using the Wrapped Website

#### Registration and content tailoring

A total of 115 participants were randomised to the intervention group and received an email link to register and access the Wrapped website. Of these, 34 (29.6%) did not follow the link to the website and two (1.7%) followed the link and registered but did not complete the tailoring items required to gain access to the site. Session recordings made by Matomo of these two individuals’ experience of completing tailoring indicated that one experienced a technical error on the website whilst the other simply stopped engaging with the content. A flow diagram showing the movement of participants through the stages of registration is provided in figure 1 of Supplementary File 3.

The 11 participants in the qualitative study that accessed the website described the process of registration as uncomplicated and similar in experience to that of other websites. Of the remaining four, one participant was disinterested in accessing the website, and the other three did not remember receiving the link and suggested sending this by SMS as well as email in the future.

> *“So I honestly must have just not completely read through it. Yeah, if I had, I probably would have gone on to the website." (PP20)*

The tailoring items on the website were intended to ensure users only received components of the intervention where they had an expressed need, a strategy intended to optimise cost-effectiveness and user engagement. As described in the method section above, participant allocation to components made by the tailoring items was compared to allocation made using the same participant responses to baseline survey items. A total of 41 participants (51.9%) were assigned all six components. As shown in Table 3 below, discrepancy existed across all components but was particularly pronounced for components two and five.

**Table 3.** Comparison of component allocation using responses to website and survey questions.

| Component | Number of participants allocated to component based on tailoring items | Number of participants with evidence of need for this component based on survey items | Discrepancy between intervention tailoring and baseline survey |
| --- | --- | --- | --- |
| 2. Condom Ordering Service | 70 | 29 | 41 (58.6%) |
| 3. Condom Carrier | 63 | 55 | 8 (12.7%) |
| 4. Condom Demo Video | 70 | 58 | 12 (17.1%) |
| 5. Discussing Condoms Videos | 59 | 44 | 15 (25.4%) |
| 6. Real Life Videos | 66 | 54 | 12 (18.2%) |

The UX review highlighted an important question; may the tone of messaging ahead of the tailoring items be inadvertently encouraging users to endorse some or all items? Messaging included “caution: you won’t be able to change your answers” and “this is the last opportunity to change your answers before they are locked in”. It was hypothesised that this alerted users of the potential to “miss out” on content unless they answered “yes” to everything, which may explain why just over half of registered users were assigned all six components. The messaging was deliberately cautionary as users were unable to return to these tailoring items to correct an error made.

#### Wrapped Website Design and Ease of Use

After completing registration and the tailoring items, participants were taken to the home page which gave them access to their assigned components, each displayed as a clickable icon. Most participants (n=71; 89.8%) accessed at least one of their assigned components, however eight participants (10.1%) bounced off the website before accessing any of the content.

Of the 71 registered users who completed the month 3 survey, the majority of participants (n=48; 67.6%) described the website as very or extremely useful and 55 participants (77.5%) rated the design of the website as good or very good. Further, participants in the qualitative study described the website as easy to navigate and use, for example, when ordering products. Though most of these participants liked the look and appearance of the website, describing it as straightforward and smart, and appreciating it’s non-medicalised feel, a minority thought the design lacked visual appeal.

> *"Yeah, it was very - it was quite smart, but it was quite bold as well and the information wasn’t overwhelming” (PP8)*

> *"I really liked it, and it was really simple to use. I liked how it wasn’t overloaded with information, and the information you wanted to find was quite simple. I liked the little icons and the style of it all, and that was really nice as well." (PP23)*

> *"It’s quite neutral colours and quite friendly. It’s not like boring, this is medical. If you go on the NHS website kind of thing, it’s all like blue and medical and that kind of thing. Whereas it was a bit more friendly, and stuff." (PP6)*

> *"Honestly it’s nothing special but it works. It’s simple and easy to navigate." (PP52)*

The UX review highlighted the problematic information architecture of the intervention: the way the content was structured on the website meant that it fell short of UX best practice, restricting navigability and opportunities for exploration. Specifically, because the tailoring items presented a unique combination of components to each participant, the components themselves were, by nature, siloed on the website, with no connecting links between them to allow deeper and more meaningful discovery of content. This lack of connecting links between content fails to conform to natural mental models that web users have; for example, after watching a video on how to put on a condom (component 4), users might naturally gravitate towards ordering condoms (component 2), but even if users have access to both, with no crosslinks made between them, this type of behaviour might not be observed. The siloed presentation of content was also recognised as limiting the potential of the homepage as a “shop window”, drawing users in through interesting presentation of content; this may have contributed to the observed 10% bounce rate off the website.

### Experience of Intervention Components

#### Condom Sample Pack

All registered participants (n=79) were assigned the Condom Sample Pack. Of these, 65 participants viewed the Condom Sample Pack page, 49 began to place an order (i.e., they began to enter information on the ordering page), and 48 (60.8%) achieved the predefined goal by completing the order. See Figure 2 of Supplementary File 3 for more details.

Of the 79 registered participants, 73 (92.4%) completed the month 12 survey. Of these, 42 (57.5%) reported that they had received the sample pack, with 38 (90.5%) reporting that they had tried at least one of the condoms, 33 (78.6%) reporting that they had tried at least one of the sachets of lubricant, 38 (90.5%) reporting that they had looked at the leaflet sent with the pack, and 26 (61.9%) reporting that they were using the sample pack box to store condoms in (its intended use).

Eight participants in the qualitative study had ordered the sample pack. These individuals described having been motivated to order the sample pack due to barriers they had previously experienced in obtaining condoms (cost, availability, and embarrassment purchasing in-store) and to facilitate experimentation with different types of condoms in order to discover their preferred type(s).

Participants reported being happy with the contents, with some pleasantly surprised by the amount and variety of condoms. The variety of condoms exceeded expectations as some participants held beliefs that free condoms would be of poor quality; participants appreciated that the condoms included in the sample pack were easy to obtain elsewhere at affordable prices.

> *I’ve always thought of it as the boy’s job. Because it was there and available, and obviously free, there was a reason to have them on me, just in case. Which is definitely the better option, than relying on somebody else perhaps. So, yeah, that was quite useful.” (PP35)*

> *"And also when the condoms arrived, it was exciting for myself and my partner. Because we were like, ooh, there’s so many different ones and we can try all of them, so that was also quite promising as well." (PP23)*

> *"It was quite good, actually, and I was actually quite surprised. I was expecting a few, and not… About one or two or three maybe, but not the variety that actually comes with it.” (PP13)*

> *“I like how it was that was the range that was given, and it’s easily accessible, and it’s not super exclusive.” (PP23)*

#### Condom Ordering Service

A total of 70 participants (88.6%) were assigned the Condom Ordering Service. Of these, 51 (72.9%) visited the Condom Ordering Service page, 38 (54.3%) proceeded with the ordering process, and 37 (52.9%) achieved the predetermined goal of placing at least one order. Twenty-five (67.6%) returned to the site on at least one subsequent occasion to place a further order. See Figure 3, Supplementary File 3 for more details. A total of 113 individual orders were placed, ranging from 1-10 orders per participant over the 12-month period, with an average of 3 orders per participant.

Of the 73 registered users who completed the month 12 survey, 64 were assigned the Condom Ordering Service. Twenty-two of these (34.4%) reported that they had placed at least one order, of which 11 (50%) had reportedly used most or all the condoms from their most recent order.

In the qualitative study, 11 participants had been assigned the Condom Ordering Service, of whom nine placed at least one order. The number of orders placed ranged from one to eight, with all but one of these participants placing more than one order. The Condom Ordering Service was well liked by participants, describing it as being discreet. Participants appreciated the variety of condoms and lubricants available to order and liked how information was provided for each condom to help them make their selection.

> *"It was pretty good, I can’t fault it for a free service." (PP52)*

> *"So it’s absolutely amazing the fact that there’s such a selection as well. And, yeah, tried both the lubes as well and it’s just great products there, so why would we not use them?’ (PP16)*

> *“Well, it’s more private and secure [than in-person settings]." (PP13)*

> *“I guess some people are a bit, I don’t know, iffy about buying them themselves, I don’t know. So I just thought, oh, actually, it’s kind of like a private way of getting this stuff, I thought from that side of it, it was quite cool.” (SP10)*

#### Condom Carrier

The Condom Carrier was assigned to 63 participants (79.7%), 49 of whom visited the Condom Carrier page, 33 proceeded with the ordering process, and 31 (49.2%) achieved the predetermined goal of placing an order. See Figure 4, Supplementary File 3 for more details.

Of the 73 registered users who completed the month 12 survey, 59 (80.8%) were assigned the Condom Carrier component, 24 (32.8%) of whom indicated they had ordered a carrier. There was a discrepancy between the order history and participants’ memory of ordering a carrier at month 12; six participants’ order history indicated they had ordered a carrier, but at M12 they reported they had not. Of the 24 who reported they had ordered a carrier, 10 (41.7%) said they had used the carrier to carry condoms, 4 (16.7%) said they had used the carrier but not for condoms, and 10 (41.7%) reported that they had not used the condom carrier at all. Reasons provided for this included not liking the colour or design, or that the carrier was not discreet enough. Further qualitative comments from the survey indicated that barriers to using the carrier included forgetting to take the carrier or condoms out, not wanting to carry something else for condoms, and feeling like it was *“Not as sexy, getting it out of a carrier takes extra time.”*

The condom carrier received mixed feedback from the qualitative participants; 10 of those interviewed had been assigned the carrier, eight of whom placed an order. Half of these participants liked the discreet and multi-purpose functionality of the carrier and thought that the quality and style was good. The other half acknowledged that although they liked the thinking behind the idea, they didn’t use the carrier as they disliked the quality and did not find it useful.

> *“So it was like discreet, and I could just have it around with me. And it’s also really useful, and I use it to store my cards in and everything like that, so it’s useful just to have it in there." (PP23)*

> *"I wasn’t a massive fan, to be honest. I just put it straight in the drawer and haven’t used it. I know it’s supposed to be inconspicuous, but it’s a bit boring. I thought it would draw more attention to itself, and it was just not the most attractive thing to look at." (PP6)*

Furthermore, the UX review questioned if the carrier itself was a reflection of participants’ actual needs and expectations, that is, whether the carrier eliminated a barrier they were experiencing and fulfilled their expectations regarding the design.

#### Condom Demo, Discussing Condoms, and Real Life Videos

A total of 70 participants (88.6%) were assigned the Condom Demo Video; 22 participants visited the related page, and 7 (10%) achieved the predetermined goal of playing the video, with two participants watching it in full. The median time spent watching the video was 165 seconds (the video was 238 seconds in length). See Figure 5, Supplementary File 3 for more details.

The Discussing Condoms Videos component was assigned to 59 participants (74.7%); 12 participants (20.3%) visited the related page and 4 (6.8%) achieved the predetermined goal of playing at least one of the seven videos. Only three of the videos were watched (any duration), with one video watched in full by one participant. For more details, including the median viewing times for each video, see Figure 6, Supplementary File 3.

The Real Life Videos component was assigned to 66 participants (83.5%); 25 participants (37.9%) visited the related page and 9 (13.6%) achieved the predetermined goal of playing at least one of the three videos. None of these were watched in full; for more details, including the median viewing time for each video, see Figure 7, Supplementary File 3.

Only half of the 10 participants in the qualitative study who had been assigned a video-related component watched any of them. Participants who did not engage with the videos described how they didn’t realise there were videos on the website or that they decided not to watch them because the titles of the videos suggested they were either not relevant to them or would be an uncomfortable watch.

> *"Other than potentially bad acting and second-hand embarrassment not really [interested in watching video] - but I imagine they would be useful for someone less informed about sex." (PP52)*

The participants who watched the Condom Demo and Discussing Condoms videos felt that they were relatable, enjoyable, and the content was well written; they also expressed a preference for the information being presented in this video format over written content. Two participants thought that the target audience was likely younger, less sexually experienced people. Although one participant thought the videos were too long, others thought the length was appropriate.

> *“It was very well thought out and scripted, and you could tell that everyone was trying to be as honest as they could…about everything. And it was almost similar to a conversation I would have with my friends.” (PP4)*

> *“Potentially, I think it’s slightly aimed at younger people, but, yeah, I thought it was really interesting. And, yeah, I would quite like my sister, for example, who is a few years younger than me, to have seen that, growing up." (PP6-aged 24)*

Most participants stated that they either did not watch the Real Life videos or couldn’t remember if they had. One participant did watch these videos and described them as relatable and interesting as well as educational.

> *"But it was definitely something I looked through, because I was like, oh, this is really interesting. And it’s not - it was real people, and it’s not porn and it’s more for actual educational purposes, and I quite liked that. It wasn’t awkward, like watching the videos you watched in school, where you cringe a bit, because your teacher is there, and it just… Yeah, I felt comfortable to watch them." (PP23)*

> *"Yeah, I definitely think I could, because it was like, well, they’re just normal people. They’re not people you think that are in porn, and they’re like naked, unrealistic and stuff. And the fact that it’s just kind of this could be an actual real-life situation, instead of the weird fantasies that I don’t really understand." (PP23)*

The UX review highlighted several issues with the video related components, such as poor descriptions of these components on the homepage as well as the descriptions for each individual video; this could lead to few individuals expecting to find them useful or relatable. The review also highlighted that the videos themselves were long, with no subtitles or transcripts, and dissimilar to video content usually seen on social media, e.g. short form content, vertical aspect ratio, etc.

## Discussion

This paper describes how participants engaged with the Wrapped intervention during an fRCT that was conducted to support planning for a definitive trial. An important aspect of this included identifying whether aspects of the intervention needed optimising to maximise user engagement, a metric understood to moderate intervention efficacy. In line with recommendations (6,7,10,26–28), assessment of engagement included examination of both online and offline behaviours and of the cognitive and affective experience. Overall, the results showed that most participants registered for the Wrapped website, found the experience of registration easy and straightforward, and the website itself easy to use. Most participants assigned to the Sample Pack and Condom Ordering Service components achieved the predetermined goal of placing an order. High rates of participants reported using the condoms they ordered and that they enjoyed the products themselves. This contrasted with the Condom Carrier component, with less than half of participants assigned this achieving the predetermined intervention goal of placing an order and low rates of reported usage; participants had mixed feelings about the carrier itself, with some finding it handy and useful for discreetly carrying condoms and others finding it unattractive and boring. Finally, very few participants assigned to the video components achieved the predetermined goal of watching any video for any length of time; participants who watched the videos found them relatable and interesting, while those who did not, either did not know the videos existed on the website or anticipated feeling awkward and chose not to watch them.

The triangulation of data from analytics, user history, survey, and qualitative data, and the UX review, helped build a more complete picture of engagement than would have been possible using a single-methods approach; using mixed methods gives meaning beyond analytics of how users interacted with the website (28) and provides a stronger rationale for optimisation of the website. Largely, the findings from each data source corroborated one another. For example, analytics data showed that most participants followed the email link to the website, registered, and completed the tailoring items without issue; this has typically been a barrier to engagement with digital interventions in previous research (29). Furthermore, participants in the surveys and qualitative interviews described the registration process and navigation of the website as easy and that the website was very or extremely useful. Perceived ease of use and perceived usefulness are important concepts to the use of digital interventions and may impact on participants’ intention to use them (30,31). Findings however did not always corroborate, and highlighted important aspects of use that might otherwise have gone unnoticed. For example, discrepancies in the allocation of intervention components based on responses to the tailoring items versus to the baseline survey indicated that participants may have been answering “yes” to all tailoring items in order not to miss out on any intervention content; a hypothesis that was further supported by the UX review. To add to this picture, several participants in the qualitative interviews said they didn’t even know the videos existed on the website. These findings highlight the problematic information architecture of the intervention website, with the tailoring items themselves a catalyst for the siloed nature of the components. Based on this, the decision was made to remove content tailoring in an optimised version of the intervention website. Breaking free from this siloed design will allow users to take a more active role in their experience of the website which may act to reduce intervention attrition (32). This change will also enable Wrapped to have more visual appeal and interest, an aspect noted as lacking by some users.

The intervention components that offered free products (Condom Sample Pack, Condom Ordering Service, and Condom Carrier) had the highest rates of predetermined goal achievement. Offline usage of the Sample Pack and Condom Ordering Service (i.e. using the condoms and reading the accompanying printed material) was also high. Participants were enthusiastic about the products they ordered from the website, with the Sample Pack and Condom Ordering Service being especially well liked for the variety of products available and its discreet and private nature. However, of the free products on the website, the Condom Carrier had lower rates of predetermined goal achievement and actual usage. Analytics data indicated that although there were occasions where participants visited the product related components and did not complete an order, the overwhelming majority did, suggesting few, if any, technical barriers to achieving the product related goals. The survey and qualitative data, however, highlighted that several participants had forgotten that they had even ordered the condom carrier 12 months after the order was placed, potentially indicating an immemorable experience. Additionally, participants had less interest in the product itself as well as less motivation to use it - barriers that analytics alone could not have identified.

The video related components had the lowest levels of predetermined goal achievement, despite qualitative feedback indicating that for those that did view them, the videos themselves were relatable and enjoyable. The UX review highlighted how the videos themselves did not conform to participants’ expectations of typical online video content (i.e., long runtime, no subtitles); this may explain why very few of the videos were watched in their entirety. Participants in the qualitative study who did not watch the videos at all commented that they anticipated they would not meet their needs. This finding might be explained by the discrepancy between expressed need as indicated at the baseline survey and the tailoring items: up to 25% of participants allocated to the video components did not have an expressed need for this content, according to their responses in the baseline survey. These components may have been engaged with less because some participants did not have a need to watch them in the first place; participants may be less likely to engage with an intervention if it does not meet their needs or expectations. Lack of engagement has been found to be indicative of lack of participant need (33,34).

Adopting mixed methodology as standard for studying engagement is recommended to fully capture the participant experience of an intervention (28). Analytics alone cannot answer why participants do or do not access certain content on the website, how the intervention is used offline, or the non-behavioural aspects of engagement. This study demonstrates how examining engagement beyond analytics allows researchers to build a more complete picture of how and why participants are using an intervention, as well as to demonstrate that analytics alone can lead researchers to incorrect conclusions. There are multiple examples outlined above of how looking beyond analytics completed our understanding of engagement: for example, understanding the impact of problematic information architecture on intervention engagement (siloed content, created by the tailoring items, contributed to participants not fully exploring and or engaging with the components), understanding why the carrier had fewer goal completions (survey and qualitative data identified it was less desirable), understanding why the videos weren’t clicked on (qualitative data identifying that users didn’t find the content or expected it to be uncomfortable to engage with).

Intervention effectiveness can only be achieved if participants engage with the intervention itself (35). The ease of using analytics to measure engagement (7) has likely contributed to the fallacy that more engagement, as measured by increased logins and time spent on the intervention, leads to more effective outcomes (35), as well as to the infrequent reporting of mixed methods of engagement (7,8). However, more engagement does not necessarily lead to better outcomes (36) - more time spent on the intervention website could indicate more technical challenges, more frustration, and more searching and scrolling for the answers that users are looking for (28). The cognitive and affective experience of the intervention itself may be as important, or even more important, than engagement as defined by analytics in determining effectiveness (37), but more research is needed to explore this (26,27,37).

Researchers are encouraged to consider the adoption of a mixed methods approach by following the example outlined in this paper, e.g. by planning meaningful measurement of analytics, user history, survey, and qualitative data, and synthesising across data sources. Examining the intervention website from a UX perspective is also recommended. This approach can lead to new insights that highlight exactly how the intervention was used online and offline as well as how participants experienced the intervention itself; the participants in the current study discussed the joy and excitement, as well as the awkwardness and embarrassment, they felt while engaging with the intervention. This depth of insight can guide researchers when optimising digital interventions.

### Limitations

Limitations of this study include the assumption that survey responses were a more accurate indication of expressed need than the tailoring items. This may not have been case thus partially removing the justification for taking out the tailoring element in the next iteration of Wrapped. Whilst this is the case, other indicators supported its removal (e.g. improvements to the information architecture). Additionally, all participants were offered a £5 voucher to register for the intervention website, which may have artificially increased the rate of registration; previous research indicates that participants with higher financial motivations may be more likely to engage with a digital intervention if there are financial incentives (38). The intervention was part of a fRCT and therefore engagement may not be reflective of actual uptake of the intervention outside of a research context. Furthermore, the fRCT was conducted during a time of fluctuating social restrictions due to the COVID-19 pandemic, which may have had an influence on participants’ engagement with Wrapped and the relevance of content to them.

### Conclusion

In conclusion, this paper demonstrates that taking a holistic view of engagement and employing a mixed methods approach can help to provide a full picture of how and why participants engage with an intervention. It has fed directly into decisions around optimisation of the Wrapped intervention (e.g. the removal of the tailoring items) as well as supported the design of future research to further explore the needs and expectations of participants (including a UX think aloud study to gather further insight into why engagement was poor with the videos, and qualitative interviews to develop personas for the Wrapped intervention). The purpose of this further research will be to maximise the future potential of the Wrapped intervention, which is key to its effectiveness, and will be tested in an upcoming large-scale RCT.

## Data Availability

The data supporting the findings of this study are publicly available from the University of Hertfordshire Research Archive (UHRA) at https://uhra.herts.ac.uk.

## Author Contributions

**Conceptualization:** Lauren Schumacher, Kayleigh Kwah, Rik Crutzen, Katherine Brown, Stephen Bremner, Louise J Jackson, Katie Newby

**Data Curation:** Lauren Schumacher, Kayleigh Kwah, Katie Newby

**Formal Analysis:** Lauren Schumacher, Kayleigh Kwah, Katherine Brown, Katie Newby

**Funding Acquisition:** Rik Crutzen, Katherine Brown, Stephen Bremner, Louise J Jackson, Katie Newby

**Investigation:** Lauren Schumacher, Kayleigh Kwah, Rik Crutzen, Katie Newby

**Methodology:** Lauren Schumacher, Kayleigh Kwah, Rik Crutzen, Katie Newby

**Project Administration:** Lauren Schumacher, Kayleigh Kwah, Katie Newby

**Supervision:** Katie Newby

**Writing – Original Draft Preparation:** Lauren Schumacher, Kayleigh Kwah, Katie Newby

**Writing – Review & Editing:** Lauren Schumacher, Kayleigh Kwah, Rik Crutzen, Katherine Brown, Stephen Bremner, Louise J Jackson, Katie Newby

## Data Availability Statement

The data supporting the findings of this study are publicly available from the University of Hertfordshire Research Archive (UHRA) at https://uhra.herts.ac.uk. The UHRA is the publicly accessible institutional repository of the University of Hertfordshire and is registered with the Registry of Research Data Repositories (re3data.org).

## Supporting Information

S1 Expressed Need and Component Assignment

S2 Survey and Tailoring Items

S3 Flowcharts

